# Importance and frequency of using esophageal pressure monitoring during ventilatory support. A cross-sectional study

**DOI:** 10.64898/2026.04.30.26352166

**Authors:** María L Giménez, Emilio Steinberg, Luis I Garegnani

## Abstract

**Background:** Esophageal pressure (P_es_) measurement has been used successfully over the past half-century to delineate the respiratory system’s physiology and mechanics. However, there is no information about the importance of P_es_ monitoring in different scenarios.

We aimed to assess the importance and frequency of P_es_ monitoring in different scenarios according to health professionals and its importance in decision-making.

**Method:** Cross-sectional study with an international survey. We included healthcare professionals dedicated to patients receiving invasive and non-invasive ventilation without limits of age, gender, experience and seniority in the position or country of residence. We used non-probabilistic snowball sampling.

**Results:** We included 152 participants, with 54.61% (83) males. The response rate to the survey questions ranged from 100% to 71.71%. Of the included participants, 91/139 (65.47%) were respiratory therapists, and 31/139 (22.30%) were Physicians. Most participants worked in mixed ICU. 109/121 (90.08%) participants considered P_es_ monitoring very important or extremely important for teaching or research. Only 32/112 (28.57%) reported using P_es_ frequently for these proposals. 49/109 (40.50%) participants considered P_es_ monitoring very important or extremely important during non-invasive ventilatory support. Only 17/112 (15.18%) reported using P_es_ frequently for these proposals. Regarding MV individualisation in ARDS during total ventilatory support, 94/121 (77.69%) participants considered P_es_ monitoring very important or extremely important. Only 33/112 (29.46%) reported using P_es_ frequently in this scenario. 90/121 (74.38%) also considered it very important or extremely important for MV individualisation in obese patients without ARDS, and 108/121 (89.26%) considered it very important or extremely important for MV individualisation in obese patients with ARDS during total ventilatory support. Only 25/112 (22.32%) and 39/112 (34.82%) reported using P_es_ frequently in these scenarios, respectively.

**Conclusions:** P_es_ monitoring was considered very important or extremely important for most assessed scenarios. Conversely, most participants rarely or never used it, although it changed therapeutic decisions often when implemented.

## Introduction

Invasive mechanical ventilation (MV) is one of the most widely used life saving techniques in the intensive care unit (ICU)(1,2). In this practice, as well as with the entire approach to critically ill patients, it is essential to consider the implementation of therapies that have an impact on the days of MV, such as the use of neuromuscular blockers, prone position or early spontaneous breathing trials (SBT)(3). Monitoring, both quantitative and qualitative, has a vital role in the management of patients with MV. Critical care patients are highly complex, therefore, adding information from measurements made at the bedside is essential for diagnosis, treatment and decision-making process(4).

Measurement of esophageal pressure (P_es_) as a surrogate for pleural pressure has been used with great success over the past half-century to delineate the physiology and mechanical properties of the respiratory system(5,6). From a scientific point of view, there is abundant evidence that P_es_ is used. Its usefulness has been well documented in ARDS, obese and obese patients with ARDS during total(7–10) and partial support(11,12), and weaning from MV. (13,14)

However, despite the evidence supporting its use, implementation in clinical practice remains limited. An epidemiological study observed that less than 1% of patients with ARDS were treated with P_es_.(15)

Furthermore, there needs to be more information about the importance of its use in different scenarios according to healthcare professionals, the frequency of its use in different clinical situations, or its importance in the decision-making process. Therefore, we conducted the following cross-sectional study to assess the importance of P_es_ monitoring in different scenarios according to health professionals, the frequency of its use in different clinical situations, and its importance in the decision-making process.

We also aimed to describe the characteristics of the professionals, the units where P_es_ monitoring is carried out, and the equipment available for P_es_ monitoring.

## Methods

This is a cross-sectional study compliant with the STrengthening the Reporting of Observational studies in Epidemiology (STROBE) checklist (16). We included healthcare professionals dedicated to patients receiving invasive and non-invasive ventilation (NIV), including physicians, respiratory therapists, nurses and any other related professionals without limits of age, gender, experience and seniority in the position or country of residence. We used non-probabilistic snowball sampling.

We collected descriptive information regarding participants’ age, gender, country, profession and years of experience working in the field. We also collected information about the institution type (academic/non-academic), setting (private institution/ public institution or social security institution), type of intensive care unit where participants performed their activities, and the educational means for acquiring P_es_ monitoring knowledge and practice. We used a 5-point Likert scale for rating the importance of P_es_ monitoring and the frequency of P_es_ monitoring in different settings, ranging from teaching, research, NIV, invasive ventilation individualisation settings in patients with ARDS, expiratory flow obstruction like asthma or COPD, obesity/overweight, abdominal hypertension or post-surgery recovery.

For data collection, we conducted anonymous multilanguage surveys using SurveyMonkey. First, we drafted a survey and circulated it for consultation among experts and professionals in critical care and MV to test content validity (See **Appendix**). The experts judged that the questionnaire items were adequate and the items were sufficient to measure the domain of interest. We did not test for construct validity due to the lack of preexisting instruments that measure similar constructs. We pilot-tested the survey for feasibility and clarity of the questions in a group of researchers unrelated to the project and incorporated their suggested changes in the final version. Between March and October 2023, we sent the survey via email, social networks and other available means of communication to authors of studies related to MV and esophageal pressure monitoring, who were invited to share the survey with other professionals.

We reported continuous variables as means and standard deviations or medians and interquartile ranges according to the distribution, analysed by visual inspection of histograms, standardised normal probability plots and the Shapiro-Wilk test. We reported categorical variables as proportions. For the statistical analysis, we used STATA 14.0 software (StataCorp LLC, USA)(17).

We conducted this research following the fundamental ethical principles of the Belmont Report, the Declaration of Helsinki, and the Nuremberg Code. The confidentiality of the data and the protection of the participants’ privacy were guaranteed, so they could not be identified.

Participation in this study was voluntary. The information provided by participants was confidential, anonymous, used for research purposes only, coded to preserve their identity, held for the duration of the investigation and destroyed at the end. The protocol for this research was approved by the Institutional Review Board (Project Number 0025-22).

## Results

One hundred and fifty-five participants accessed the survey via email and social networks. Three participants did not consent to the study and were excluded from the analyses. We finally included 152 participants, of which 35 (23.03%) accessed the survey in English and 117 (76.97%) in Spanish. The response rate to the survey questions ranged from 100% to 71.71%. See the participants’ characteristics in **Table 1**.

**Table 1.**
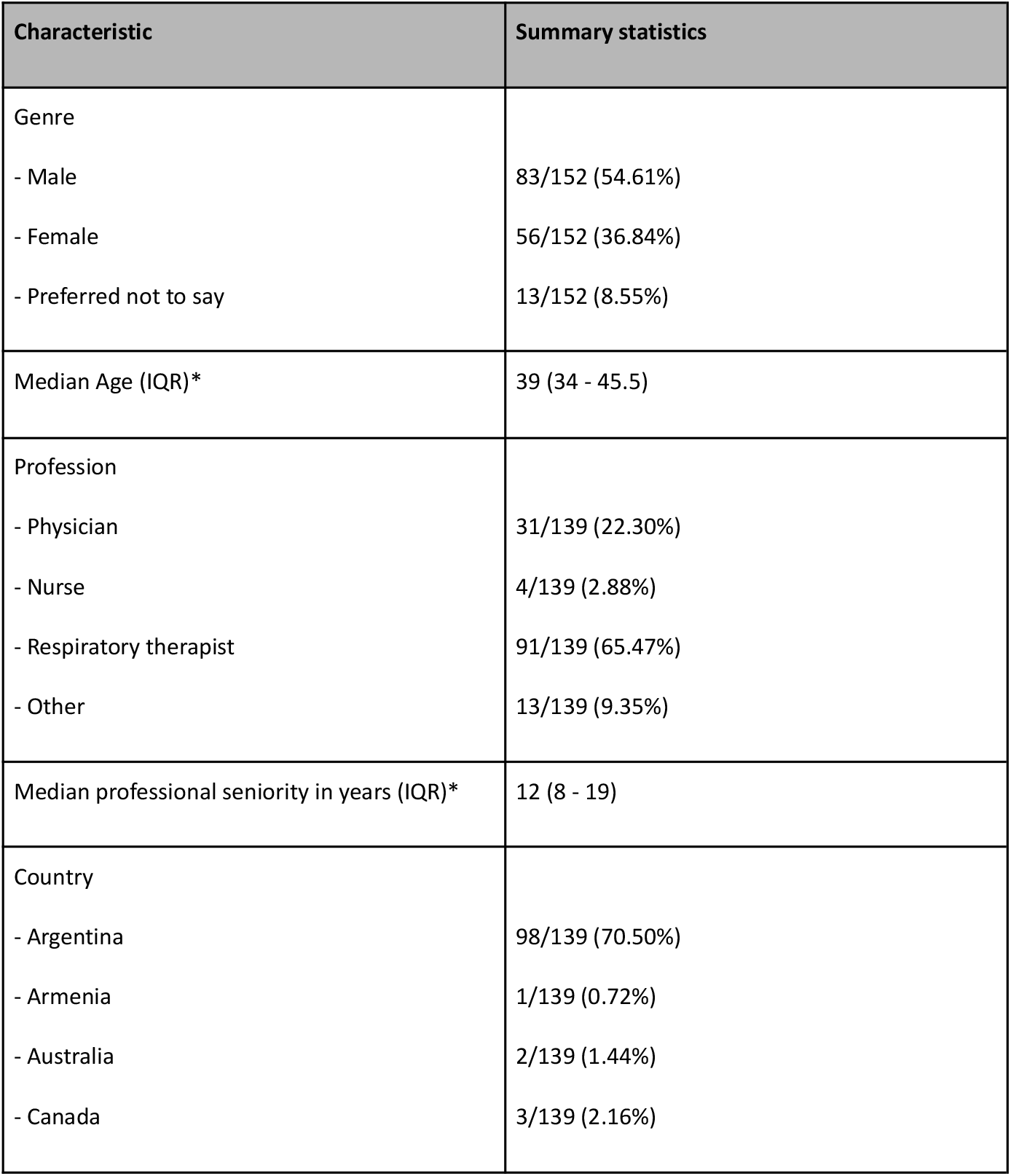
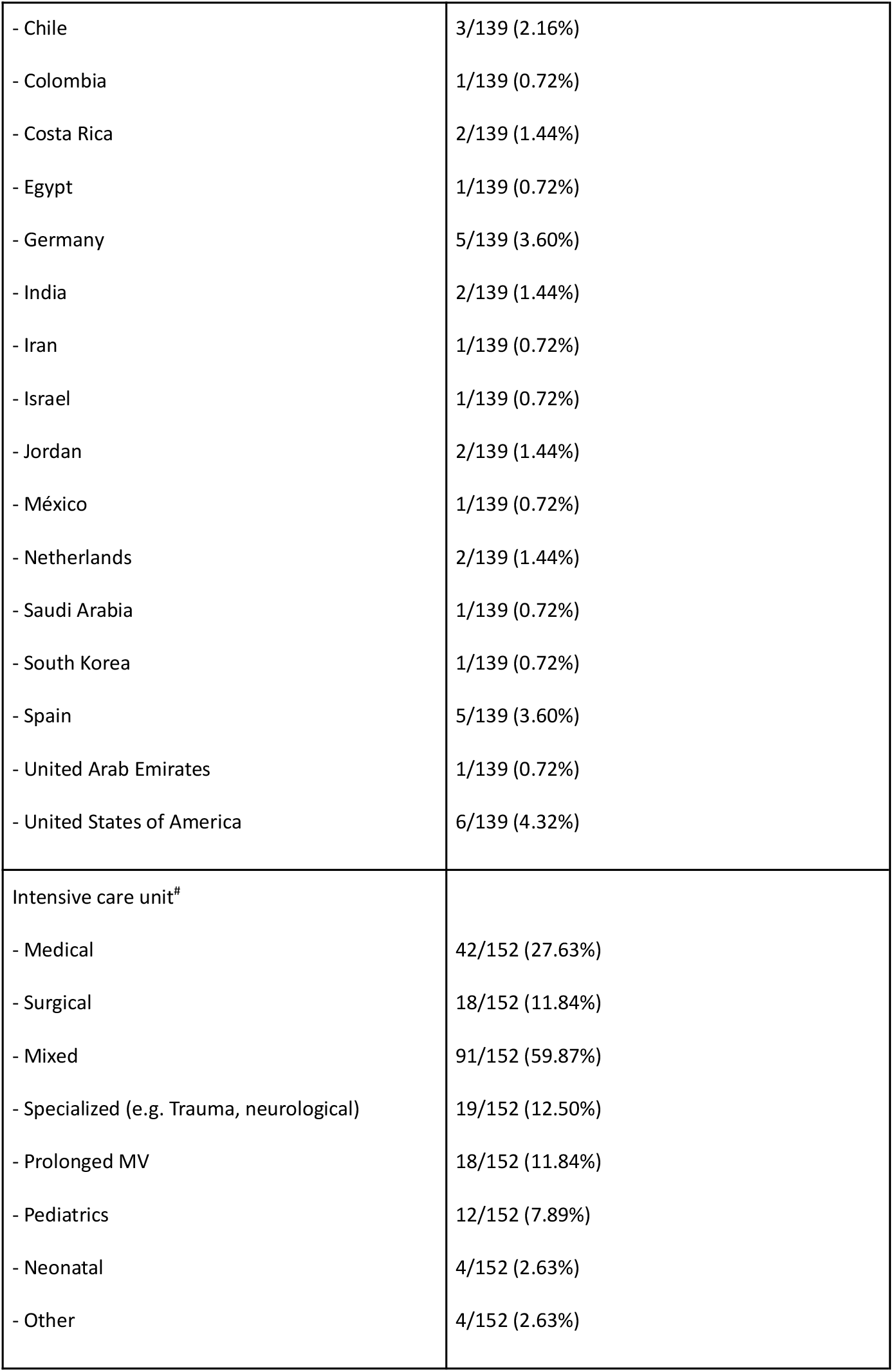
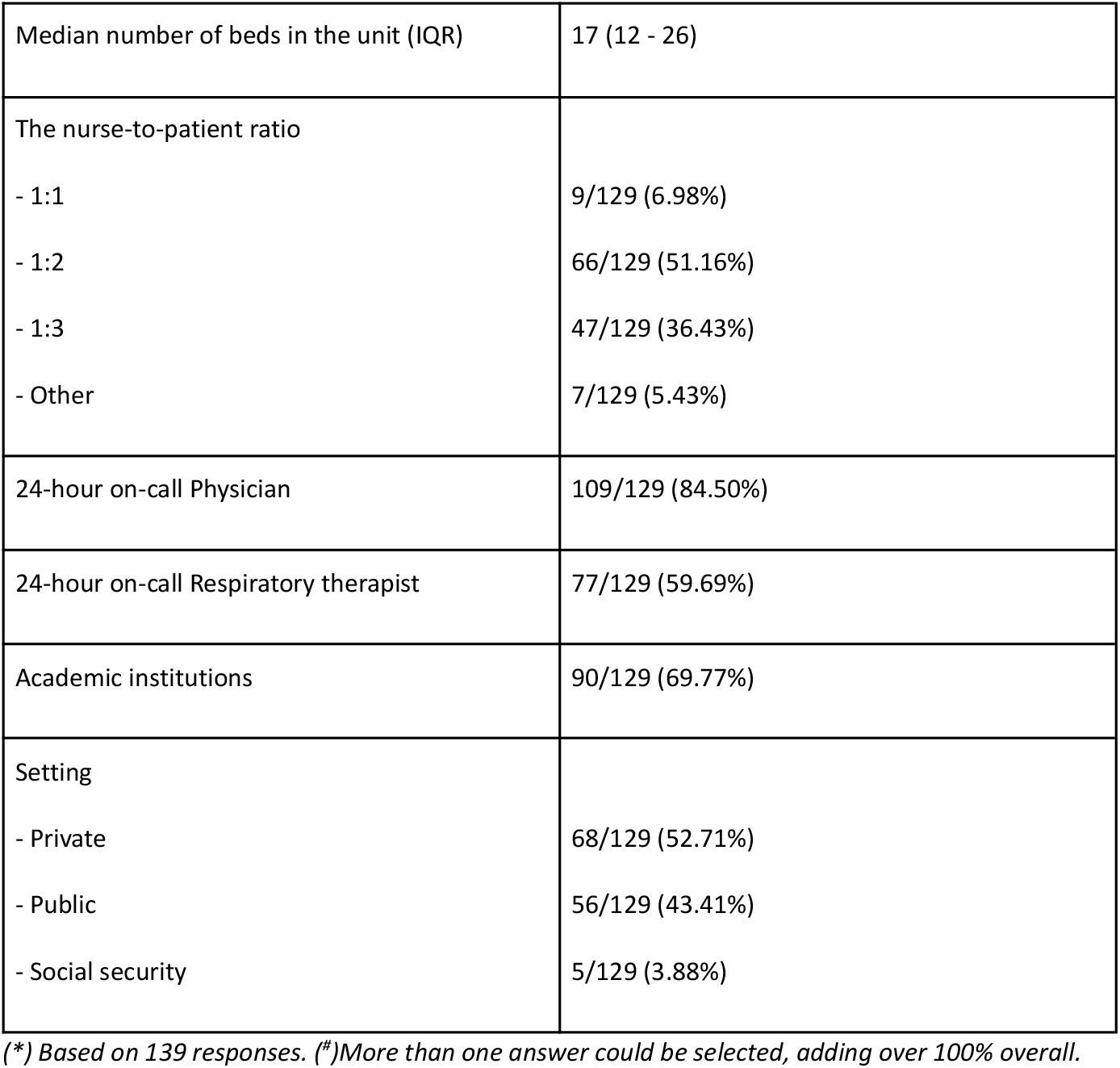
Participant’s characteristics.

Regarding the means for acquiring P_es_ monitoring knowledge and practice, 46/139 participants (33.09%) reported being guided by mentors, 17/139 participants (12.23%) reported studying introductory courses, 16/139 participants (11.51%) reported studying P_es_ monitoring by their initiative, 10/139 participants (7.19%) reported learning through workshops, 8/129 (5.76%) reported other means for learning P_es_ monitoring (most of them related to workplaces facilities and residencies) and 4/129 (2.88%) reported being part of fellow programs or academic scholarships program in advanced monitoring. 38/139 participants (27.34%) reported not having completed any actual learning program related to P_es_ monitoring.

About the importance of P_es_ monitoring in different scenarios, 109/121 (90.08%) participants considered it very important or extremely important for teaching or research proposes. Only 32/112 (28.57%) reported using P_es_ frequently for these proposes. 49/109 (40.50%) participants considered P_es_ monitoring very important or extremely important during high flow oxygen therapy (HFOT) or NIV. Only 17/112 (15.18%) reported using P_es_ frequently for these proposes. Regarding MV individualisation in ARDS during total ventilatory support, 94/121 (77.69%) participants considered P_es_ monitoring very important or extremely important. Only 33/112 (29.46%) reported using P_es_ frequently in this scenario. 90/121 (74.38%) also considered it very important or extremely important for MV individualisation in obese patients without ARDS, and 108/121 (89.26%) considered it very important or extremely important for MV individualisation in obese patients with ARDS during total ventilatory support. Only 25/112 (22.32%) and 39/112 (34.82%) reported using P_es_ frequently in these scenarios, respectively. P_es_ monitoring was considered very important or extremely important for MV individualisation in the post-operative by 47/121 (38.84%) participants, for MV individualisation during total ventilatory support in patients with abdominal hypertension by 91/121 (75.21%) participants and for setting MV in patients with airflow obstruction by 68/121 (56.20%) participants. Only 15/112 (13.39%), 22/112 (19.64%), and 16/112 (14.29%) reported using P_es_ frequently in these scenarios, respectively. See a detailed description of the importance of P_es_ monitoring during total ventilatory support in **Figure 1**. See a detailed description of the frequency of using P_es_ in **Table 2**.

**Table 2.** Frequency of using esophageal pressure monitoring in different scenarios.

| Scenarios | Summary statistics |  |  |  |  |
| --- | --- | --- | --- | --- | --- |
|  | Never | Rarely | Sometimes | Often | Very often |
| **Total ventilatory support** |  |  |  |  |  |
| Teaching/research | 25/112<br>(22.32%) | 20/112<br>(17.86%) | 35/112<br>(31.25%) | 20/112<br>(17.86%) | 12/112<br>(10.71%) |
| Non-invasive ventilatory support | 69/112<br>(61.61%) | 16/112<br>(14.29%) | 10/112<br>(8.93%) | 10/112<br>(8.93%) | 7/112<br>(6.25%) |
| MV individualisation in ARDS | 28/112<br>(25.00%) | 21/112<br>(18.75%) | 30/112<br>(26.79%) | 24/112<br>(21.43%) | 9/112<br>(8.04%) |
| MV individualisation in the obese without ARDS | 37/112<br>(33.04%) | 22/112<br>(19.64%) | 28/112<br>(25%) | 18/112<br>(16.07%) | 7/112<br>(6.25%) |
| MV individualisation in the obese with ARDS | 28/112<br>(25.00%) | 21/112<br>(18.75%) | 24/112<br>(21.43%) | 25/112<br>(22.32%) | 14/112<br>(12.50%) |
| MV individualisation in patients with airflow obstruction | 52/112<br>(46.43%) | 26/112<br>(23.21%) | 18/112<br>(16.07%) | 12/112<br>(10.71%) | 4/112<br>(3.57%) |
| **Partial ventilatory support** |  |  |  |  |  |
| Asynchrony detection | 50/112<br>(44.64%) | 23/112<br>(20.54%) | 20/112<br>(17.86%) | 13/112<br>(11.61%) | 6/112<br>(5.35%) |
| Assistance titration | 50/112<br>(44.64%) | 22/112<br>(19.64%) | 22/112<br>(19.64%) | 11/112<br>(9.82%) | 7/112<br>(6.25%) |
| Respiratory muscles assessment | 55/112<br>(49.11%) | 23/112<br>(20.54%) | 19/112<br>(16.96%) | 11/112<br>(9.82%) | 4/112<br>(3.57%) |
| Weaning assessment | 55/112<br>(49.11%) | 24/112<br>(21.43%) | 15/112<br>(13.39%) | 13/112<br>(11.61%) | 5/112<br>(4.46%) |

*Pes: Esophageal pressure. MV: Mechanical ventilation. ARDS: Acute Respiratory Distress Syndrome.*

**Figure 1.**
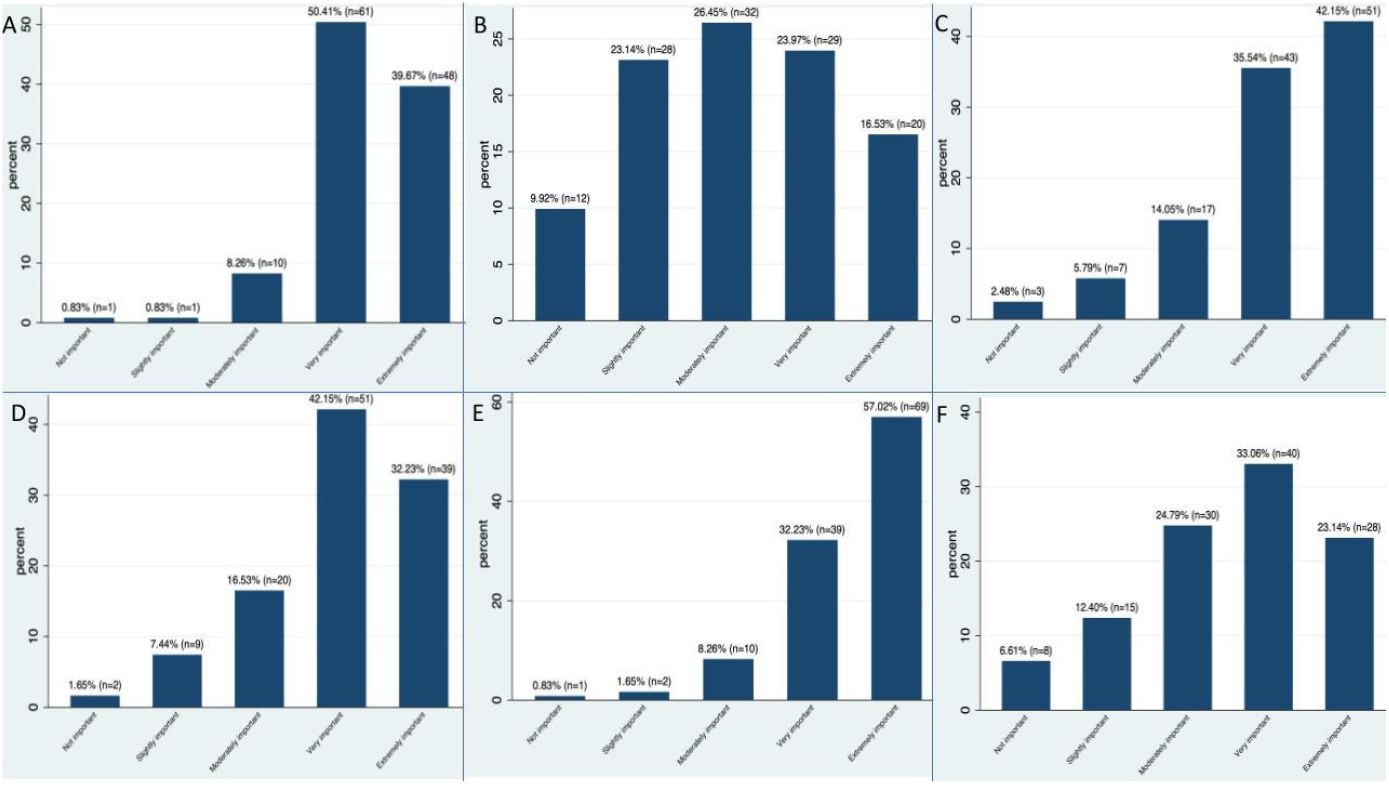
The importance of using esophageal pressure monitoring during total ventilatory support in different scenarios. A: Teaching/research; B: Non-invasive ventilatory support; C: Mechanical ventilation individualisation in ARDS during total ventilatory support; D: Mechanical ventilation individualisation in the obese without ARDS during total ventilatory support; E: Mechanical ventilation individualisation in the obese with ARDS during total ventilatory support; F: Mechanical ventilation individualisation in patients with airflow obstruction during total ventilatory support

During partial ventilatory support, P_es_ monitoring was considered very important or extremely important for asynchrony detection, assistance titration, respiratory muscle assessment and weaning assessment by 82/121 (67.77%), 78/121 (64.46%), 78/121 (64.46%) and 73/121 (60.33%) respectively. Only 19/112 (16.96%), 18/112 (16.07%), 15/112 (13.39%), and 18/112 (16.07%) reported using P_es_ frequently in these scenarios, respectively. See a detailed description of the importance of P_es_ monitoring during partial ventilatory support in **Figure 2**. See a detailed description of the frequency of using P_es_ in **Table 2**.

**Figure 2.**
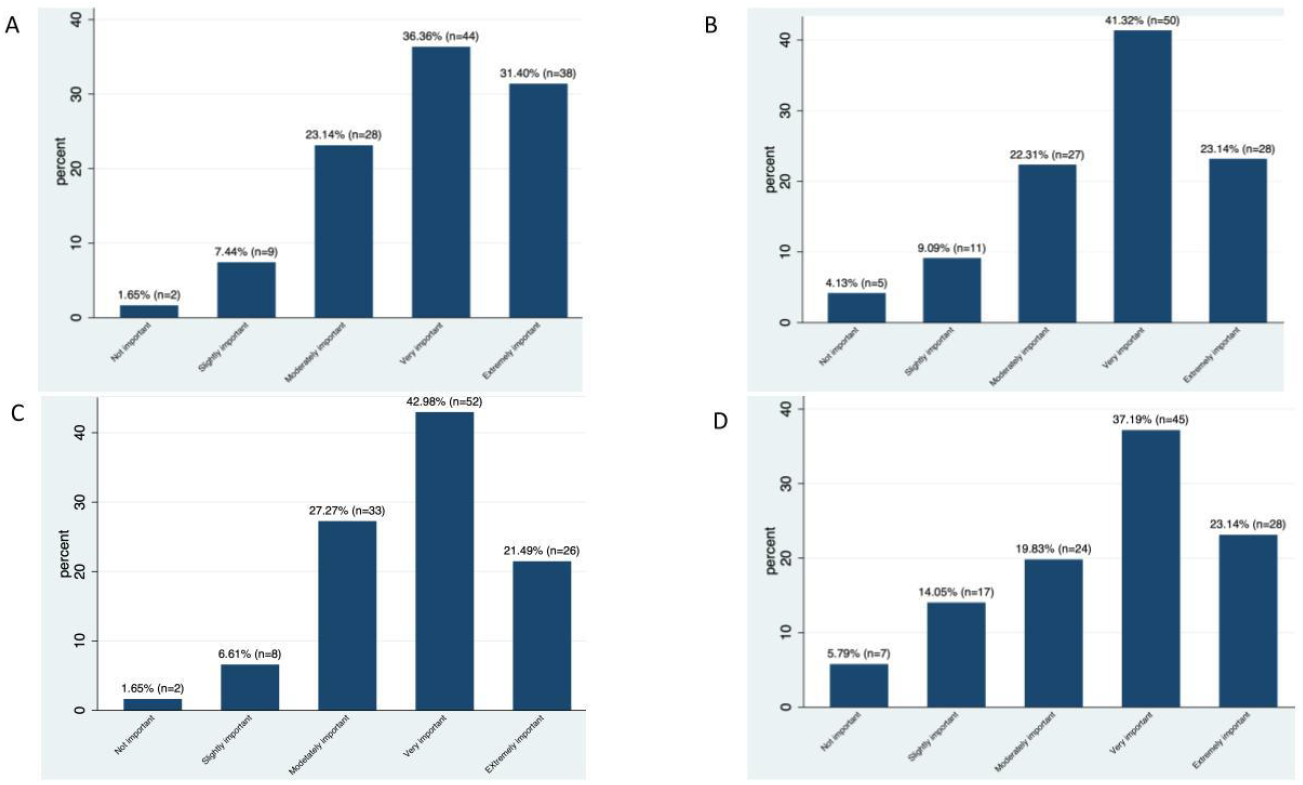
The importance of using esophageal pressure monitoring during partial ventilatory support in different scenarios. A: Asynchrony detection during partial ventilatory support; B: Assistance titration during particle ventilatory support; C: Respiratory muscles assessment during partial ventilatory support; D: Weaning assessment during partial ventilatory support.

The information obtained through P_es_ monitoring changed the therapeutic decisions in 79/111 (71.17%) participants. The equipment used for P_es_ monitoring was highly variable, ranging from 46/109 (42.2%) participants using specific P_es_ monitors, 17/109 (15.6%) participants using mechanical ventilators with onboard P_es_ monitoring, 8/109 (7.34%) participants using multiparametric monitors and 6/109 (5.5%) participants using analogue manovacuometers. 31/109 (28.44%) participants referred to not having any P_es_ monitoring devices available in their units.

## Discussion

We included primarily young men with many years of experience in their field. Most included participants were respiratory therapists working in mixed ICUs with many available beds and a sizeable nurse-to-patient ratio in the private sector within academic institutions. Most participants acquired P_es_ monitoring knowledge with a mentor’s guidance, although a remarkable proportion reported not having completed any learning program. P_es_ monitoring was considered very important or extremely important for most assessed scenarios, except for its use during NIV. Conversely, most participants rarely or never used P_es_ monitoring for the assessed scenarios, although it changed therapeutic decisions often when implemented.

In 2016, the first epidemiological study including patients with ARDS requiring MV reported using P_es_ monitoring in 19 of 3022 patients, which represents 0.8%.(15) In our survey, we found a higher reported use of P_es_ monitoring than that of Bellani et al. Nevertheless, the reported use may be considered low in relation to the importance this tool represents to the participants. Scarce equipment availability, lack of healthcare personnel training and primarily limited scientific evidence supporting P_es_ monitoring use could justify its rare application in patients with ARDS. At this juncture, recent ARDS treatment guidelines do not recommend its use in PEEP titration.(3)(18) Recently, Wisse et al, described the main use of P_es_ for PEEP titration and measuring lung and chest wall compliance(19). This would be extremely important, especially in patients with ARDS, as it would help quantify the impact of chest mechanics on the respiratory system (20) and thus allow for the individualized setting of MV.(21)

EPvent 1 and EPVent2 studies found no improvements in mortality or number of days free from MV compared to conventional PEEP titration strategies (PEEP/FIO2 Table). (7)(8)This is why experts refer to P_es_-guided PEEP as a method that requires further studies to ensure that end-expiratory pressure does not cause overdistension in non-dependent areas. Without these data, the best method to individualize PEEP in clinical practice remains unclear.(3)(18)

Pathophysiology of obese patients is interesting in terms of the relationship between the chest wall, lungs, and abdomen. Physiological studies began using P_es_ as early as 1985, finding that end-expiratory P_es_ was higher than atmospheric pressure, favored by the weight of the chest wall and increased intra-abdominal volume. Failing to consider this aspect when setting MV in obese patients may lead to underestimation of PEEP, resulting in impaired gas exchange and increased risk of atelectrauma.(22) P_es_ guided MV to assess transpulmonary pressure is essential for individualizing MV settings in obese patients. (22,23) In 2016, Pirrone et al. showed that clinicians commonly used PEEP levels that were inadequate and insufficient in morbidly obese patients.(22,23) Rowley et al. reported similar data in 2021, finding improvements in respiratory system compliance (C) and increased PaFiO_2_ once patients were ventilated and ventilator parameters were guided by P_es_ monitoring.(24)

The results of this survey have once again emphasized the importance of P_es_ monitoring in obese populations, but its usage was around 20%, with a higher rate in obese patients with ARDS. However, there are no studies demonstrating improvements in mortality, days of MV, or reduction in ICU length of stay associated with the use of P_es_ in obese patients.

The evidence regarding the use of P_es_ during SBT is scarce and not entirely conclusive regarding the mechanical determinants of failure.(25)(26) The best approach to perform an SBT is yet controversial,(27) (28) which may be one of the reasons explaining the considerable variability in this practice.(29) However, not all methods are physiologically similar.(30)(31) In our study, 50% of the participants reported never implementing P_es_ during SBT, while 20% said they had almost never done so.

Approximately 55% to 65% of patients who initiate weaning are successfully extubated on the first attempt.(32,33) It is likely that this population does not require extensive monitoring complexity. However, the remaining 45 to 35% could benefit from a more precise approach in terms of monitoring both muscular effort and respiratory system mechanics, (34) making P_es_ a potentially valuable asset.

Monitoring effort, respiratory mechanics, and patient-ventilator synchrony during spontaneous MV remains challenging. (11) Approximately 70% of respondents do not use P_es_ monitoring during partial support. This contrasts with the findings reported by Wisse et al.(19), where more than 50% of respondents reported using it in this scenario. A possible explanation for this discrepancy is that most respondents in Wisse et al.’s study were from high-income countries, whereas our participants were primarily from middle-income countries. (35)(36) In fact, the cost of using P_es_, along with the availability of materials and equipment, seems to be one of the barriers limiting its routine use.(19) The availability and simplicity of non-invasive effort monitoring tools (37)(38)(39) and respiratory mechanics (40) may explain the limited implementation of P_es_ monitoring. However, expiratory effort could limit the accuracy of both muscular and mechanical assessments. (41) (42) In a scenario of potential risk for developing patient self-inflicted lung injury, (43) coupled with the fact that patients who regain muscular contraction after a period of sedation and fail the transition to spontaneous ventilation have worse outcomes, (44) the implementation of P_es_ could prove beneficial.

One of the main and overarching goals, both in terms of devices and scenarios of NIV, is the reduction of work of breathing (WOB). (45,46) In this regard, CPAP and bilevel NIV have shown effects on WOB in both hypoxemic (47)(48)(47,49)(50) and hypercapnic respiratory failure. (51) (52)

Approximately 75% of participants in our study reported never or rarely implementing P_es_ monitoring in non-invasive support as reported by Wisse et al.(19) Potential technical challenges, coupled with the lack of recommendations in clinical practice guidelines, could justify the low frequency of use.(53,54) However, published studies in recent years may help guide the way to increase its implementation in the future. (49)(47)(55)

Healthcare facilities often combine teaching and research under the scope of academic activity when describing their tasks. However, these are two very distinct fields. On one hand, research disseminates new knowledge, and education prepares us to apply it. In our survey, around 50% of respondents stated that they use P_es_ for ‘teaching and research’ purposes, a value that is not reflected in the number of publications in the region. Therefore, we assume that most professionals use P_es_ for educational purposes while also employing it in patient care. In this sense, integrating P_es_ teaching during clinical education at the bedside emerges as a strategy that facilitates the understanding of complex respiratory pathophysiological processes, which are difficult to grasp when taught separately from clinical practice. P_es_ monitoring allows us to correlate pathophysiological processes with clinical findings and ventilatory strategies directly and in real-time. Therefore, implementing programs aimed at trainees using P_es_ could be beneficial.

Our study is not free from limitations. First, we intended to include participants worldwide, disseminating the survey through different communications channels. Nevertheless, we included mainly participants from South America, which may limit the generalizability of our findings to other geographic regions and the majority of the surveyors were respiratory therapists. Second, our survey may have been too long for the participants, as the response rate diminished from the first questions to the last ones, reflecting a possible exhausting effect on the participants. Nevertheless, the overall response rate was acceptable and aligned with other international surveys(56). Although the topic may be highly relevant to clinical practice, the survey does not address the crucial aspects of identifying barriers and potential solutions. Finally, given the novelty of our approach, we were unable to test for construct validity, although we received feedback from experts in the field and pilot-tested the survey before collecting data, following standard methods.

## Conclusions

P_es_ monitoring was considered very important or extremely important for most assessed scenarios, except for its use during NIV. Conversely, most participants rarely or never used P_es_ monitoring for it, although it changed therapeutic decisions often when implemented.

P_es_ monitoring may serve for assessing the different respiratory system components behaviours. Nevertheless, P_es_ monitoring availability and use in specific scenarios remains limited in relation to the importance clinician gives to it. P_es_ monitoring use remains almost exclusive for MV individualisation in ARDS, obesity and research or teaching.

## Glossary

Pes: Esophageal pressure
MV: mechanical ventilation (MV
SBT: spontaneous breathing trials (SBT
ICU: intensive care unit (ICU)
NIV: non-invasive ventilation (NIV)
ARDS: acute respiratory distress sindrome
COPD: cronic obstructive pulmonary disease
HFOT: high flow oxygen therapy
WOB: work of breathing
CPAP: continuous positive airway pressure
PEEP: positive end respiratory presure

## Declaration of generative AI in scientific writing

all the authors declare not using AI in this paper.

## Data sharing statement

The data supporting this study’s findings are available from the corresponding author upon reasonable request.

