## Appendix for "Importance and frequency of using esophageal pressure monitoring during ventilatory support. A cross-sectional study"

**Use of esophageal pressure (Pes) monitoring during ventilatory support**

**Information before you start**

*This survey is anonymous and voluntary and aims to know the characteristics of the professionals and units using esophageal pressure monitoring (Pes) and determine the setting and pathologies in which professionals consider it important. By completing and submitting your answers, you are authorizing us to collect the information you provide. This information will be completely confidential, anonymous and used only for research purposes. Your data will be encrypted to preserve your identity, preserved for the duration of the investigation and destroyed at the end of it*

**Do you want to participate?**

Yes

No

This question requires an answer.

**Section 1: Respondent Characteristics**

**What is your gender?**

Female

Male

Other

**How old are you? (in years)**

The comment you entered is not in a valid format.

This question requires an answer.

**What is your profession?**

Physician

Nurse

Physical/Respiratory Therapist

Please enter a comment.

Other (please specify)

**What is your professional seniority? (in years)**

This question requires an answer.

**How did you learn to use and apply esophageal pressure (Pes) monitoring?**

Courses

Workshops

Fellows

Guided by mentors

Personal initiative

I have no training in esophageal pressure monitoring.

Please enter a comment.

Other (please specify)

This question requires an answer.

**In which country do you work?**

Afghanistan

Albania

Algeria

Andorra

Angola

Antigua and Barbuda

Argentina

Armenia

Australia

Austria

Azerbaijan

Bahamas

Bahrain

Bangladesh

Barbados

Belarus

Belgium

Belize

Benin

Bhutan

Bolivia

Bosnia and Herzegovina

Botswana

Brazil

Brunei Darussalam

Bulgaria

Burkina Faso

Burundi

Cape Verde

Cambodia

Cameroon

Canada

Central African Republic

Republic of Chad

Chile

China

Colombia

Comoros

Congo

Costa Rica

Ivory Coast

Croatia

Cuba

Cyprus

Czech Republic

Democratic People's Republic of Korea

Democratic Republic of Congo

Denmark

Djibouti

Dominica

Dominican Republic

Ecuador

Egypt

El Salvador

Equatorial Guinea

Eritrea

Estonia

Ethiopia

Fiji

Finland

France

Gabon

Gambia

Georgia

Germany

Ghana

Greece

Grenade

Guatemala

Guinea

Guinea-Bissau

Guyana

Haiti

Holy See

Honduras

Hungary

Iceland

India

Indonesia

Iran

Irak

Ireland

Israel

Italy

Jamaica

Japan

Jordan

Kazakhstan

Kenya

Kiribati

Kuwait

Kyrgyzstan

Laos (Lao People's Democratic Republic)

Latvia

Lebanon

Lesotho

Liberia

Libya

Liechtenstein

Lithuania

Luxembourg

Madagascar

Malawi

Malaysia

Maldives

Mali

Malt

Marshall Islands

Mauritania

Mauricio

Mexico

Micronesia (Federated States of)

Monaco

Mongolia

Montenegro

Morocco

Mozambique

Myanmar

Namibia

Nauru

Nepal

Netherlands

New Zealand

Nicaragua

Níger

Nigeria

Norway

Oman

Pakistan

Palau

Panama

Papua New Guinea

Paraguay

Peru

Philippines

Poland

Portugal

Qatar

Republic of Korea

Republic of Moldova

Rumania

Russian Federation

Rwanda

Saint Kitts and Nevis

St. Lucia

St. Vincent and the Grenadines

Samoa

San Marino

Sao Tome and Principe

Saudi Arabia

Senegal

Serbia

Seychelles

Sierra Leone

Singapur

Slovakia

Slovenia

Solomon Islands

Somalia

South Africa

South Sudan

Spain

Sri Lanka

State of Palestine

Sudan

Suriname

Swaziland

Sweden

Switzerland

Syrian Arab Republic

Tajikistan

Thailand

North Macedonia

East Timor

Togo

Tonga

Trinidad and Tobago

Tunisia

Turkey

Turkmenistan

Tuvalu

Uganda

Ukraine

United Arab Emirates

United Kingdom of Great Britain and Northern Ireland

United Republic of Tanzania

United States of America

Uruguay

Uzbekistan

Vanuatu

Venezuela

Vietnam

Yemen

Zambia

Zimbabwe

This question requires an answer.

**Section 2: Characteristics of the unit where the respondent works**

**Indicate the type of unit where you work (you can select more than one option)**

Medical intensive care unit

Surgical intensive care unit

Mixed intensive care unit

Specialized surgical intensive care unit (for example trauma, neurological, cardiovascular, etc.)

Long-term mechanical ventilation facilities

Pediatric intensive care unit

Neonatal intensive care unit

Please enter a comment.

Other (please specify)

This question requires an answer.

**Indicate the number of available beds in your unit**

The comment you entered is not in a valid format.

This question requires an answer.

**Is there an onsite critical medical care specialist 24 hours a day, 7 days a week?**

Yes

No

This question requires an answer.

**Is there an onsite specialist Respiratory Physiotherapist 24 hours a day, 7 days a week?**

Yes

No

This question requires an answer.

**What is the nurse-to-patient ratio of your unit?**

1:1

1:2

1:3

Other

This question requires an answer.

**What type of institution does your unit belong to?**

Academic

Non-academic

This question requires an answer.

What is the setting of your institution?

Public

Private

Social security

This question requires an answer.

**Section 3: Importance of Pes monitoring during mechanical ventilation in different settings**

*Indicate the importance you assign to Pes monitoring in the following settings/strategies. We define “total ventilatory support" as mechanical ventilation without respiratory muscular effort from the patient. We define"partial ventilatory support" as mechanical ventilation with respiratory muscle effort from the patient*

**Teaching/research**

Extremely important

Very important

Moderately important

Slightly important

Not at all important

This question requires an answer.

**Non-invasive ventilatory support (HFNC/NIV)**

Extremely important

Very important

Moderately important

Slightly important

Not at all important

This question requires an answer.

**Mechanical ventilation (MV) individualization in ARDS during total ventilatory support**

Extremely important

Very important

Moderately important

Slightly important

Not at all important

This question requires an answer.

**MV individualization in the obese WITHOUT ARDS during total ventilatory support**

Extremely important

Very important

Moderately important

Slightly important

Not at all important

This question requires an answer.

**MV individualization in the obese WITH ARDS during total ventilatory support**

Extremely important

Very important

Moderately important

Slightly important

Not at all important

This question requires an answer.

**MV individualisation in the Postoperative Period during total ventilatory support**

Extremely important

Very important

Moderately important

Slightly important

Not at all important

This question requires an answer.

**MV individualization in patients with abdominal hypertension during total ventilatory support**

Extremely important

Very important

Moderately important

Slightly important

Not at all important

This question requires an answer.

**MV individualisation in the patient with airflow obstruction during total ventilatory support**

Extremely important

Very important

Moderately important

Slightly important

Not at all important

This question requires an answer.

**Asynchrony detection during partial ventilatory support**

Extremely important

Very important

Moderately important

Slightly important

Not at all important

This question requires an answer.

**Assistance titration during partial ventilatory support**

Extremely important

Very important

Moderately important

Slightly important

Not at all important

This question requires an answer.

**Respiratory muscles assessment during partial ventilatory support**

Extremely important

Very important

Moderately important

Slightly important

Not at all important

This question requires an answer.

**Weaning assessment during partial ventilatory support**

Extremely important

Very important

Moderately important

Slightly important

Not at all important

This question requires an answer.

**Section 4: Frequency of Pes monitoring utilization during MV in different settings**

*Indicate how often you use Pes monitoring in the following settings/strategies. We define "total ventilatory support" as MV without respiratory muscle effort from the patient. We define "partial ventilatory support" as MV with respiratory muscle effort from the patient.*

**Teaching/research**

Very often

Often

Sometimes

Rarely

Never

This question requires an answer.

**Non-invasive ventilatory support (HFNC/NIV)**

Very often

Often

Sometimes

Rarely

Never

This question requires an answer.

**Mechanical ventilation (MV) individualisation in ARDS during total ventilatory support**

Very often

Often

Sometimes

Rarely

Never

This question requires an answer.

**MV individualisation in the obese WITHOUT ARDS during total ventilatory support**

Very often

Often

Sometimes

Rarely

Never

This question requires an answer.

**MV individualisation in the obese WITH ARDS during total ventilatory support**

Very often

Often

Sometimes

Rarely

Never

This question requires an answer.

**MV individualization in the Postoperative Period during total ventilatory support**

Very often

Often

Sometimes

Rarely

Never

This question requires an answer.

**MV individualisation in patients with abdominal hypertension during total ventilatory support**

Very often

Often

Sometimes

Rarely

Never

This question requires an answer.

**MV individualization in the patient with airflow obstruction during total ventilatory support**

Very often

Often

Sometimes

Rarely

Never

This question requires an answer.

**Asynchrony detection during partial ventilatory support**

Very often

Often

Sometimes

Rarely

Never

This question requires an answer.

**Assistance titration during partial ventilatory support**

Very often

Often

Sometimes

Rarely

Never

This question requires an answer.

**Respiratory muscles assessment during partial ventilatory support**

Very often

Often

Sometimes

Rarely

Never

This question requires an answer.

**Weaning assessment during partial ventilatory support**

Very often

Often

Sometimes

Rarely

Never

This question requires an answer.

**Section 5: Decisions based on Pes monitoring**

**Has the information obtained through Pes monitoring generated changes in your therapeutic decisions?**

Yes

No

This question requires an answer.

**Section 6: Available equipment for Pes monitoring**

**What equipment do you use for Pes monitoring during mechanical ventilation?**

Pes specific monitor

Mechanical ventilator with Pes monitorig

Multi parametric monitor

Manovacuometer

I do not have equipment for Pes monitoring

Please enter a comment.

Other (please specify)

This question requires an answer.

**What type of catheter do you use for Pes monitoring? (you can select more than one option)**

Commercial

Hand made

With gastric balloon

I do not have Pes monitoring catheters

Please enter a comment.

Other (please specify)

This question requires an answer.
